# Improved evaluation of left ventricular hypertrophy using the spatial QRS-T angle by electrocardiography

**DOI:** 10.1101/2022.01.10.22268661

**Authors:** Maren Maanja, Todd T Schlegel, Rebecca Kozor, Ljuba Bacharova, Timothy C. Wong, Erik B Schelbert, Martin Ugander

**Affiliations:** Department of Clinical Physiology, Karolinska University Hospital, and Karolinska Institutet, Stockholm, Sweden; Nicollier-Schlegel SARL, Trélex, Switzerland; Department of Cardiology, Royal North Shore Hospital, Sydney, Australia; Kolling Institute, Royal North Short Hospital, and Northern Clinical School, Sydney Medical School, University of Sydney, Sydney, Australia; International Laser Center, Bratislava, Slovak Republic; Institute of Pathophysiology, Medical School, Comenius University, Bratislava, Slovak Republic; Department of Medicine, University of Pittsburgh Medical Center, Pittsburgh, PA, USA

## Abstract

**Background:** Conventional electrocardiographic (ECG) signs of left ventricular hypertrophy (LVH) lack sensitivity. The aim was to identify LVH based on an abnormal spatial peaks QRS-T angle, and evaluate its diagnostic and prognostic performance compared to that of conventional ECG criteria for LVH.

**Methods:** This was an observational study with four cohorts, all with a QRS duration <120 ms: (1) Healthy volunteers to define normality (n=921), (2) Separate healthy volunteers to compare test specificity (n=461), (3) Patients with at least moderate LVH by cardiac imaging (Imaging-LVH) to compare test sensitivity (n=225), and (4) Patients referred for cardiovascular magnetic resonance imaging to evaluate the combined outcome of hospitalization for heart failure or all-cause death (Clinical-Consecutive, n=783).

**Results:** An abnormal spatial peaks QRS-T angle was defined as exceeding the upper limit of normal, which was found to be ≥ 40° for females and ≥ 55° for males. In healthy volunteers, the specificity of the QRS-T angle to detect LVH was 96% (females) and 98% (males). In Imaging-LVH, the QRS-T angle had a higher sensitivity to detect LVH than conventional ECG criteria (93–97% vs 13–56%, *p*<0.001 for all). In Clinical-Consecutive, of those who did not have any LVH, 238/556 (43%) had an abnormal QRS-T angle, suggesting it can occur even without LVH. There was an association with outcomes in univariable analysis for the QRS-T angle, Cornell voltage, QRS duration, and Cornell product (hazard ratios 1.68–2.5, p<0.01 for all) that persisted in multivariable analysis only for the QRS-T angle and QRS duration (p<0.001 for both).

**Conclusions:** An increased QRS-T angle rarely occurred in healthy volunteers, was a mainstay of moderate or greater LVH, was common in clinical patients without LVH but with cardiac co-morbidities, and associated with outcomes. Thus, an increased QRS-T angle identifies left ventricular electrical remodeling that can occur in the absence of LVH detected by imaging. The improved diagnostic and independent prognostic performance for the QRS-T angle suggests that it should be investigated when ECGs are evaluated.

## Introduction

Left ventricular (LV) hypertrophy (LVH) is a hallmark of cardiac end-organ damage often related to diabetes (1), hypertension (2), and obesity (3). It is also associated with poor cardiovascular outcomes (4). LVH is defined as an increase in LV mass and is typically diagnosed by non-invasive cardiac imaging such as echocardiography or cardiovascular magnetic resonance imaging (5). However, the increase in LV size is also a substrate for electrical remodeling (6), and here the ECG provides unique information.

ECG diagnosis of LVH is typically based on increased QRS complex amplitudes, which have a low sensitivity and high or varying specificity (7). The electrical remodeling in LVH is complex and affects cardiomyocytes as well as the extracellular matrix (8), and extends beyond changes in QRS amplitudes. Notably, both electrical and anatomical changes attributed to LVH convey independent prognostic information (9). Taken together, these findings raise several important and related issues. First, the diagnostic performance of ECG criteria for detecting anatomical changes associated with LVH is limited, and this is likely related to the inherent differences between anatomy and electrophysiology. Second, and consequently, it has been suggested that a term such as left ventricular electrical remodeling (LVER) might be more appropriate than LVH when referring to ECG changes (5, 10). Thus, it may be more important to define electrical changes as a deviation from normal ECG values measured in healthy volunteers, rather than attempting to use the ECG to identify an anatomical measure, as well as evaluating the prognostic performance of the ECG abnormality.

An abnormal vectorcardiographic QRS-T angle is not only an important sign of LVH (11), but also an independent predictor of cardiovascular and all-cause mortality (12–14). Clinical observations inspired the hypothesis that the vectorcardiographic QRS-T angle could provide diagnostic value beyond those of conventional ECG criteria for LVH. Therefore, the aims of this study were to a) propose a sex-specific upper limit of normal for the spatial peaks QRS-T angle derived from the resting 12-lead ECG, and b) investigate the diagnostic and prognostic performance of the QRS-T angle compared to conventional ECG criteria for LVH. We hypothesized that the QRS-T angle would provide diagnostic and prognostic value beyond that of conventional ECG criteria for LVH.

## Methods

### Participants and outcomes

In this observational cohort study, patients were identified retrospectively from three prospectively acquired databases divided into four cohorts: (1) healthy volunteers used to define normality and a threshold for the QRS-T angle (Healthy-Derivation cohort), (2) healthy volunteers used to evaluate and compare the specificity of all studied measures (Healthy-Validation cohort), (3) clinical patients with at least moderate LVH by cardiac imaging, to evaluate the sensitivity of all studied measures (Imaging-LVH cohort), and (4) clinical patients referred for CMR imaging who also had follow-up data on hospitalization for heart failure or all-cause death, to evaluate the diagnostic and prognostic value of all studied measures (Clinical-Consecutive cohort). Mortality status was ascertained by medical record review and Social Security Death Index queries as previously described (15). The study was approved by the respective human subject research ethics committees, and all participants provided written informed consent.

### Healthy volunteer cohorts

A database with digital ECG recordings from healthy volunteers was divided into two parts where two thirds were used for Healthy-Derivation, and one third for Healthy-Validation. The Healthy-Derivation and Healthy-Validation cohorts were included from three different sites in the USA, as previously described in detail (16). Briefly, all healthy volunteers were low risk and asymptomatic, without evidence of cardiovascular or other systemic disease based on a physical examination and negative history, and with a clinically normal conventional ECG. If LVH or hypertrophic cardiomyopathy was suspected based on clinical assessment of the ECG, an echocardiogram was performed to rule out the given suspected pathology(s), and the subject was not included in any of the Healthy cohorts. Asymptomatic volunteers who received treatment for diabetes, hypertension, or active smokers were excluded.

### Imaging-LVH cohort

The Imaging-LVH cohort was included from seven different sites in the USA, Sweden, and Venezuela, as previously described in detail (16). Briefly, inclusion criteria for the Imaging-LVH cohort were an ECG acquired within 30 days of the cardiac imaging examination. The Imaging-LVH cohort was limited to at least moderate LVH by imaging for the purposes of evaluating the sensitivity of the ECG criteria in a population with definite LVH. Exclusion criteria were a non-sinus or paced rhythm, left or right bundle branch block, pre-excitation, an incomplete ECG recording, and imaging findings of a predominant cardiac pathology other than LVH, such as myocardial infarction or extensive non-ischemic scarring by CMR, or at least moderate valvular disease by echocardiography.

### Clinical-Consecutive cohort

The Clinical-Consecutive cohort included consecutive patients referred clinically for CMR at University of Pittsburgh Medical Center, PA, USA, between 2008 and 2017, and with follow-up until April 2018. Inclusion criteria for the Clinical-consecutive cohort were an ECG with sinus rhythm and a QRS duration <120 ms, and an ECG acquired within 30 days of the CMR examination. Exclusion criteria were ECG confounders such as atrial fibrillation or flutter, abundant premature ventricular contractions (bigeminy/trigeminy), paced rhythm, hypertrophic cardiomyopathy, congenital heart disease, Takotsubo cardiomyopathy, amyloidosis, siderosis, Fabry’s disease, and poor CMR image quality.

### ECG acquisition and analysis

ECG data for the healthy volunteers and Imaging-LVH cohorts included resting ECG acquired at 1000 Hz sampling rate, with an acquisition duration of at least 10 seconds. ECG data for the Clinical-Consecutive cohort was collected from the local clinical digital ECG database (MUSE® Cardiology Information System, Version 8.0 SP2, GE Healthcare, Chicago, IL, USA) and exported into anonymized .xml files with coded subject identification. ECG data for the Clinical-Consecutive cohort included resting ECG acquired at sampling rates of 250 or 500 Hz, with an acquisition duration of 10 seconds. The following conventional ECG variables were automatically analyzed using in-house developed software (16) according to established published criteria: Sokolow-Lyon index, defined as the sum of the S wave in lead V1 (S_V1_) plus the larger of the R_V5_ or R_V6_, where LVH was defined as >3.5 mV (17); Cornell voltage, defined as S_V3_ plus R_aVL_, where LVH is defined as >2.0 mV for females and >2.8 mV for males (18); Cornell voltage product, defined as the QRS duration times the Cornell voltage, where LVH is defined as ≥244 mV·ms (19); and the QRS duration, defined as the time from the beginning of the Q-wave to the end of the S-wave, where increased QRS duration was defined as ≥100 ms (20). The vectorcardiogram was derived from the resting 12-lead ECG using the previously validated Kors’ regression method (21), and the derived spatial peaks QRS-T angle was defined as the angular difference, in three-dimensional space, between the maximum magnitude of the vectors of the QRS and T loops, respectively (22). See Figure 1 for a schematic illustration of the spatial peaks QRS-T angle, and Figure 2 for ECGs from two representative cases from the study.

**Figure 1.**
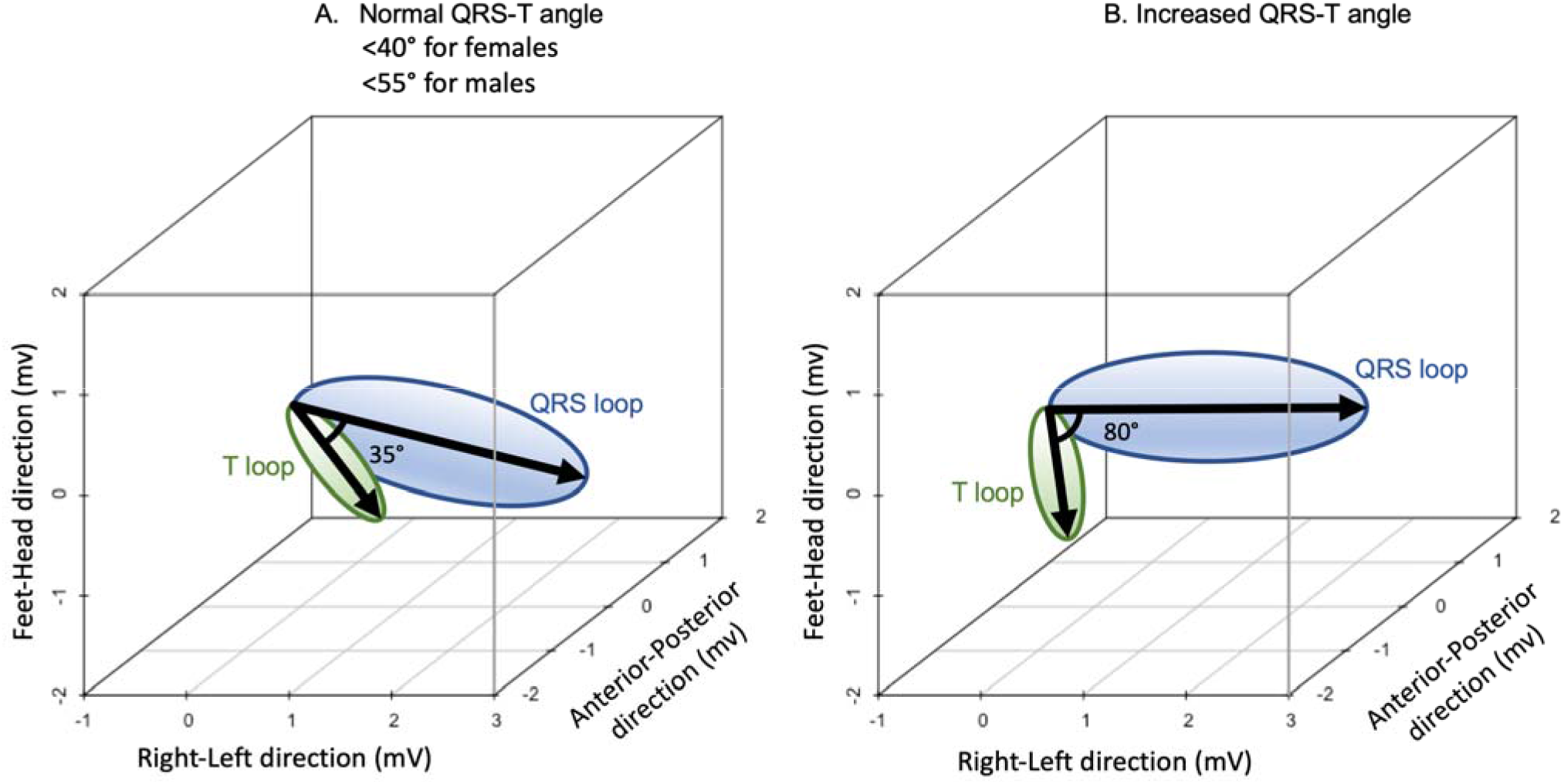
Schematic illustration of the QRS-T angle, a vectorcardiographic measure derived from a digital conventional 12-lead ECG. The vectorcardiogram represents the electrical activity in millivolts (mV) in three dimensions along the left-right, feet-head and anterior-posterior axes. The QRS loop (blue) and T loop (green) represent the direction and peak magnitude of depolarization and repolarization, respectively, of the left ventricle. The thick arrows show the largest magnitudes of the respective loops. The angle in three-dimensional space between the largest magnitude of the QRS loop and T loop is referred to as the spatial peaks QRS-T angle. **A.** Schematic example of a normal QRS-T angle of 35° in a healthy volunteer. **B.** Schematic example of an increased QRS-T angle of 80°.

**Figure 2.**
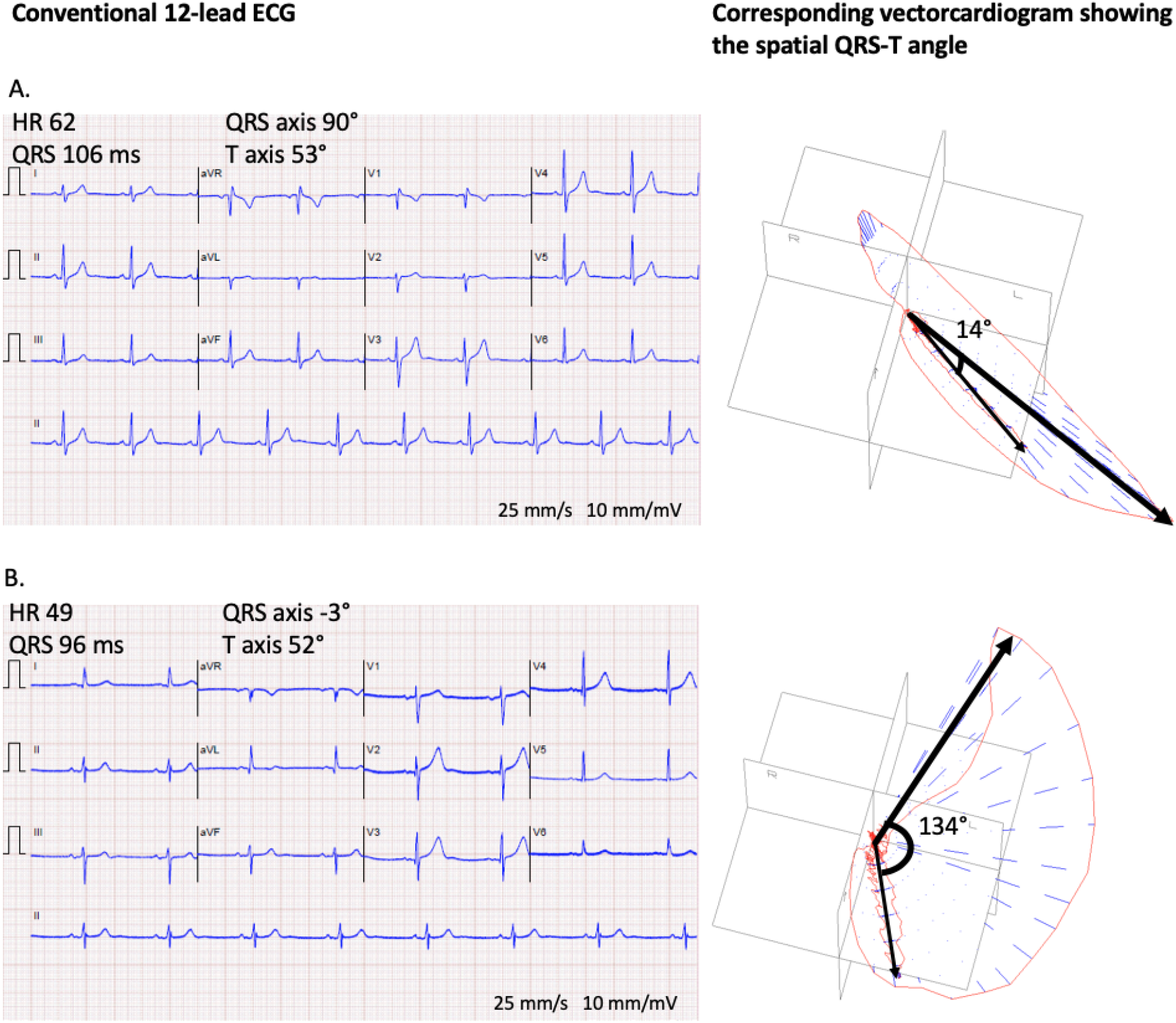
Two representative examples of different QRS-T angles from the study. **A.** A normal conventional 12-lead ECG in a healthy male in their 50s, with a corresponding derived vectorcardiographic spatial peaks QRS-T angle of 14° (normal). **B.** A normal conventional 12-lead ECG in a male in their 80s with imaging-proven LVH, with a corresponding derived spatial peaks QRS-T angle of 134° (abnormal). HR denotes heart rate. Note, the QRS axis and T axis noted in the figure are from the frontal plane, whereas the spatial peaks QRS-T angle is calculated in three-dimensional space from the derived vectorcardiogram.

### Echocardiographic and cardiovascular magnetic resonance imaging analysis

In the Imaging-LVH cohort, LVH was defined as being at least moderate according to the guidelines of the American Society of Echocardiography (23). In the Clinical-Consecutive cohort, LVH was defined as either a left ventricular mass index (LVMI) or left ventricular end diastolic volume index (LVEDVI) greater than the 95% upper limit of normal for age- and sex-matched healthy volunteers with no history of cardiac disease or known risk factors according to published normal values (24). The Clinical-Consecutive cohort underwent CMR to define measures known to associate with prognosis. These measures included left ventricular global longitudinal strain (GLS), which is a marker for impaired left ventricular function, left ventricular ejection fraction (LVEF), myocardial extracellular volume fraction (ECV), LVMI, LVEDVI, infarct size, and non-ischemic scar size. These measures were acquired and quantified according to clinical standards as previously described in detail (25).

### Statistical analysis

Statistical analysis was performed in R version 3.4.3 (R Foundation for Statistical Computing, Vienna, Austria). The sex-specific cut-off for an increased derived spatial peaks QRS-T angle was defined as the upper limit of the 95% confidence interval of normal in the Healthy-Derivation cohort. The sensitivity to detect LVH by the QRS-T angle was evaluated in the Imaging-LVH and Clinical-Consecutive cohorts, respectively. The specificity to detect LVH by the QRS-T angle was evaluated in the Healthy-Validation and Clinical-Consecutive cohorts, respectively. The area under the receiver operating characteristic curve (AUC) was evaluated in the combined Healthy-Validation and Imaging-LVH cohorts to include participants and patients with and without LVH, and Clinical-Consecutive cohort, respectively, with respect to the QRS-T angle and the conventional ECG-LVH criteria, and compared using DeLong’s test.

Using univariable Cox regression and Kaplan-Meier analysis, the QRS-T angle and conventional ECG measures for LVH were also compared in their ability to predict the combined, and individual, outcome of survival free from hospitalization for heart failure or all-cause death, ranking each according to the Wald chi-square value. Multivariable Cox regression was performed using measures that were not mathematically related to each other, and also significantly associated with outcomes in univariable Cox regression. Hazard ratios were analyzed as binary variables with cut-offs for LVH, as described above, as these cut-offs are used clinically. To investigate whether the QRS-T angle and conventional criteria correctly classified LVH in the same patients, the percentage of agreement, and Cohen’s kappa statistic were calculated. To further investigate the incremental prognostic value for the QRS-T angle, net reclassification index (NRI) was calculated in the Clinical-Consecutive cohort at one year of follow-up. NRI can be used to show the correctness of reclassification of subjects based on a new model, and calculated as (26): NRI = (number of correctly reclassified events – number of incorrectly reclassified events) + (number of correctly reclassified nonevents – number of incorrectly reclassified nonevents). The adjusted R-square (R^2^) in multivariable linear regression was used to evaluate the combined relative contributions of CMR measures to the QRS-T angle. The Kolmogorov-Smirnov test was used to test if data was normally distributed, and differences between subgroups’ baseline data was tested using the Mann-Whitney U test or chi-square test, as appropriate, and described using the median [interquartile interval], or percentage, respectively. A *p*-value <0.05 was considered statistically significant.

## Results

### Baseline characteristics

A total of 2390 patients and healthy volunteers with QRS duration <120 ms and an ECG without rhythm confounders were included in the four cohorts: Healthy-Derivation (n=921, age 34 [27–48] years, full range 20 – 85 years, 39% female), Healthy-Validation (n=461, age 35 [27–48] years, 39% female), Imaging-LVH (n=225, age 63 [53–75] years, 60% female), and Clinical-Consecutive (n=783, age 55 [43–64] years, 44% female). A total of 30 females and 28 males in the Healthy-Derivation cohort were ≥60 years old. The Healthy-Derivation and Healthy-Validation cohorts did not differ with respect to age and sex distribution. The baseline characteristics of the Clinical-Consecutive cohort are presented in Table 1. In the Clinical-Consecutive cohort, 155 patients experienced events over a follow-up period of 5.7 [4.4–6.6] years: 113 (14.4 %) deaths, 68 (8.7%) hospitalizations for heart failure, and 26 (3.3%) with both. Compared to patients with a normal QRS-T angle, patients with an increased QRS-T angle had more CMR abnormalities, including LVH, and cardiovascular co-morbidities at baseline, reflecting a greater severity or burden of cardiovascular disease.

**Table 1.** Baseline characteristics for the Clinical-Consecutive cohort.

| Characteristic | All | Normal<br>QRS-T angle | Increased<br>QRS-T angle | p-value |
| --- | --- | --- | --- | --- |
| Number, n (%) | 783 (100) | 377 (48) | 406 (52) | n/a |
| Age, y | 55 (43–64) | 53 (38–62) | 57 (46–65) | <0.001 |
| Female sex, % | 342 (44) | 149 (40) | 193 (48) | 0.02 |
| LVH | 227 (29) | 59 (18) | 168 (41) | <0.001 |
| ECV, % | 27.6 (25.3–30.3) | 26.8 (24.8–29.1) | 28.5 (25.7–31.1) | <0.001 |
| EDVI, mL/m<sup>2</sup> | 78.3 (66.1–97.3) | 74.5 (63.7–89.3) | 84.5 (67.6–111.2) | <0.001 |
| GLS, % | -16.2 (-18.8– -12.3) | -17.6 (-19.5 – -15.3) | -13.9 (-17.3 – -9.8) | <0.001 |
| LVEF, % | 58.5 (47.0–65.0) | 61.0 (55.3–66.0) | 55.0 (34.3–63.0) | <0.001 |
| LVMI, g/m<sup>2</sup> | 55.3 (44.9–67.1) | 51.7 (43.6–61.4) | 59.4 (46.8–73.8) | <0.001 |
| BMI, kg/m<sup>2</sup> | 28.6 (24.4–33.9) | 28.3 (24.4–33.5) | 29.0 (24.4–34.0) | 0.41 |
| BSA, m<sup>2</sup> | 2.0 (1.8–2.2) | 2.0 (1.8–2.2) | 2.0 (1.8–2.2) | 0.28 |
| Infarct, n (%) | 165 (21) | 43 (11) | 122 (30) | <0.001 |
| Infarct size*, % | 14.7 (5.9–22.3) | 8.8 (4.4–17.5) | 15.8 (7.7–24.1) | <0.001 |
| Non-ischemic scar, n (%) | 137 (17) | 50 (13) | 87 (21) | 0.003 |
| Non-ischemic scar size*,<br>% | 2.9 (1.5–5.7) | 2.6 (0.7–6.1) | 2.9 (1.5–5.1) | 0.50 |
| *Conventional ECG-LVH criteria* |  |  |  |  |
| Sokolow-Lyon, mV | 1.8 (1.4–2.3) | 1.7 (1.4–2.2) | 1.9 (1.4–2.1) | 0.39 |
| Cornell voltage, mV | 1.4 (1.0–1.8) | 1.2 (0.95–1.6) | 1.6 (1.2–2.1) | <0.001 |
| Cornell product, mV*ms | 132 (94–176) | 114 (82–150) | 152 (110–206) | <0.001 |
| QRS duration, ms | 90 (84–98) | 88 (82–96) | 92 (84–100) | 0.03 |
| *Comorbidity, n, (%)* |  |  |  |  |
| Hypertension | 398 (51) | 158 (42) | 240 (52) | <0.001 |
| Diabetes mellitus | 171 (22) | 57 (15) | 114 (25) | <0.001 |
| *Smoking status, n, (%)* |  |  |  |  |
| Current smoker | 129 (16) | 47 (12) | 82 (18) | 0.004 |
| Ex-smoker | 246 (31) | 112 (30) | 134 (29) | 0.32 |
| *Medication, n, (%)* |  |  |  |  |
| ACEi/ARB | 315 (40) | 114 (30) | 201 (44) | <0.001 |
| Aspirin or other<br>antiplatelets | 397 (51) | 157 (42) | 240 (52) | <0.001 |
| Beta-blockers | 387 (49) | 143 (38) | 244 (53) | <0.001 |
| Digoxin | 23 (3) | 4 (1) | 19 (5) | 0.01 |
| Insulin | 121 (15) | 40 (11) | 81 (17) | <0.001 |
| Loop diuretics | 164 (21) | 45 (12) | 119 (26) | <0.001 |
| Oral hypoglycemics | 54 (7) | 12 (3) | 42 (9) | <0.001 |
| Statins | 315 (40) | 124 (33) | 191 (42) | <0.001 |
| *General indication for CMR: known or suspected disease, n (%)* |  |  |  |  |
| Arrhythmia or syncope | 147 (19) | 59 (16) | 88 (22) | 0.03 |
| Coronary artery disease /<br>vasodilator stress test | 365 (47) | 230 (61) | 135 (33) | <0.001 |
| Dyspnea | 56 (7) | 38 (10) | 18 (4) | 0.002 |
| Non-ischemic<br>cardiomyopathy | 181 (23) | 114 (38) | 67 (17) | <0.001 |
| Pre-operation evaluation | 37 (5) | 21 (6) | 16 (4) | 0.28 |
| Valvular disease | 48 (6) | 24 (6) | 24 (6) | 0.79 |

Continuous data are given as median (interquartile interval), or number (%), and compared between the Normal QRS-T angle group and Increased QRS-T angle groups using Mann-Whitney U test or Chi-square, respectively. ACEi indicates angiotensin-converting enzyme inhibitors; ARB, angiotensin II receptor blocker; BMI, body mass index; BSA, body surface area; CMR, cardiovascular magnetic resonance imaging; ECV, extracellular volume fraction; LVEDVI, left ventricular end-diastolic volume index; GLS, global longitudinal strain; LVEF, left ventricular ejection fraction; LVMI, left ventricular mass index. \* denotes that the descriptive data are presented only for those patients with infarct or non-ischemic scar, respectively.

### Normal values for the QRS-T angle

The Healthy-Derivation cohort was used to define the upper limit of the sex-specific 95% confidence interval of normal for the derived QRS-T angle. In this cohort of healthy volunteers, the upper limit of normal for the QRS-T angle differed with regards to sex but not age (results not shown). Consequently, sex-specific normal values were presented for the whole cohort regardless of age. The mean values ±1.96 SD for the QRS-T angle were 19.7±19.9° for females and 26.6±28.3° for males. This yielded sex-specific thresholds for the QRS-T angle of ≥40° for females, and ≥55° for males.

### The ECG in imaging-verified LVH

The specificity of the QRS-T angle in the Healthy-Validation cohort was 172/180 (96%) in females and 274/281 (98%) in males. In the Imaging-LVH cohort, the sensitivity of the QRS-T angle for detecting LVH was higher than that of conventional ECG criteria (93–97% vs 13– 56%, *p*<0.001 for all, see Figure 3), illustrating that an increased QRS-T angle is present in nearly all LVH of moderate or greater severity, and to a far greater extent than positive conventional ECG criteria for LVH. In the Clinical-Consecutive cohort, the sensitivity of the QRS-T angle for detecting any LVH defined as a LVMI or a LVEDVI above the upper limit of normal by CMR was 54/77 (79%) in females and 105/150 (71%) in males, again superior to that of the conventional ECG criteria (*p*<0.001 for all). In the Clinical-Consecutive cohort, the specificity of the QRS-T angle for LVH was 133/265 (50%) in females and 185/291 (64%) in males, and was lower compared to that of the conventional criteria (*p*<0.01 for all). Of the patients in the Clinical-Consecutive cohort who did not have any LVH, 238/556 (43%) had an increased QRS-T angle.

**Figure 3.**
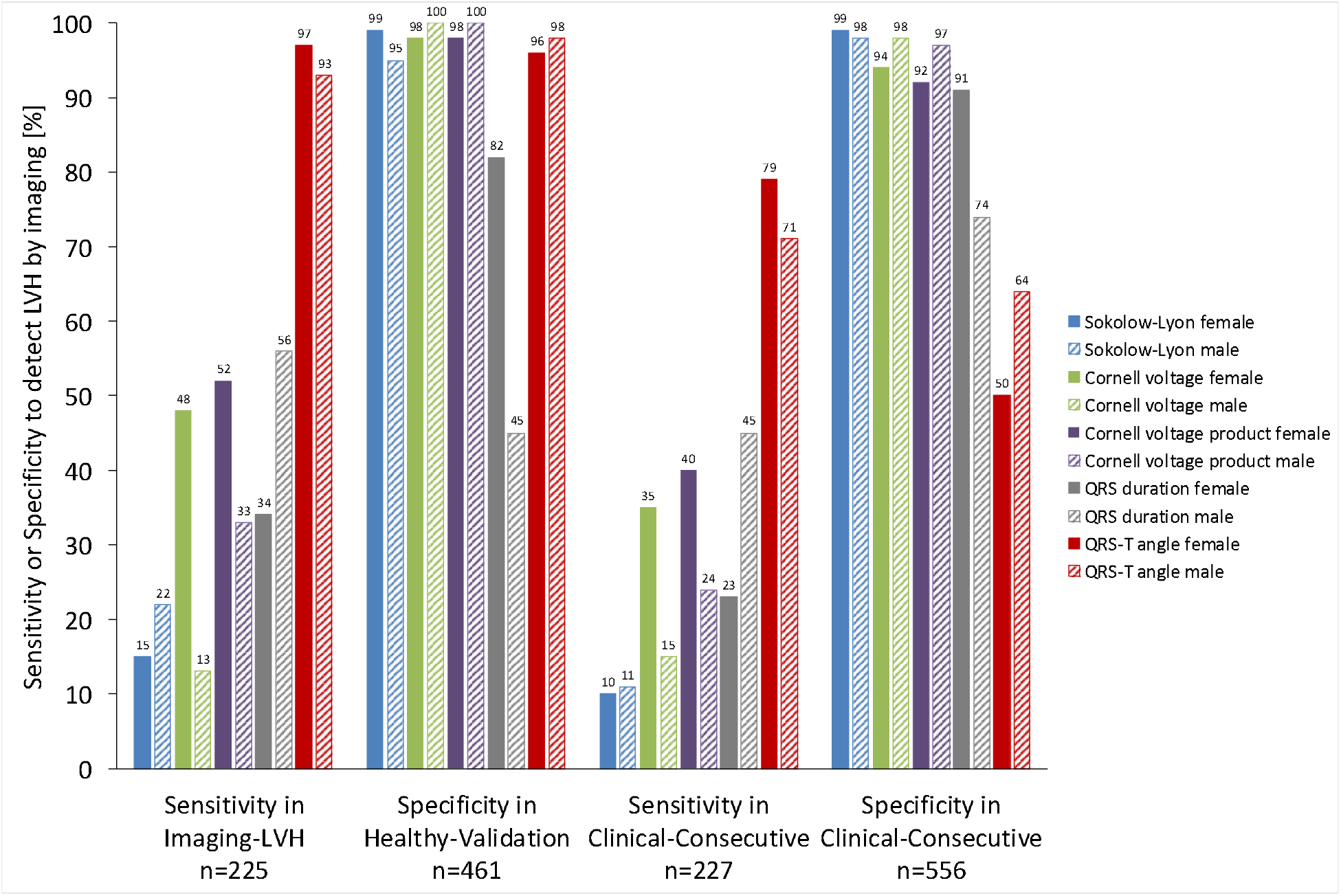
Sex-specific sensitivity or specificity for detecting anatomically defined left ventricular hypertrophy (LVH) in the respective cohorts. In both the Imaging-LVH and Clinical-Consecutive cohorts, an increased QRS-T angle had a higher sensitivity for LVH than all other ECG measures (p<0.05 for all). Among females in the Healthy-Validation cohort, the specificity of the QRS-T angle for LVH was higher than that of QRS duration (p<0.001), lower than Cornell product (p<0.05), and did not differ significantly from that of the other measures (p>0.05 for all). Among males in the Healthy-Validation cohort, the specificity of the QRS-T angle for LVH was higher than that of QRS duration (p<0.001), lower than Cornell voltage and Cornell voltage product (p<0.05 for both), and did not differ significantly from Sokolow-Lyon (p=0.12). In the Clinical-Consecutive cohort, the QRS-T angle had a lower specificity than the other ECG measures (p<0.05 for all) despite its excellent specificity in the Healthy-Validation cohort, suggesting possible electrical identification of subclinical disease in patients who do not yet fulfill imaging criteria for LVH.

In the Clinical-consecutive cohort, a total of 19 patients (8%) with LVH were detected by both the Sokolow-Lyon index and the QRS-T angle, 47 patients (21%) by both Cornell voltage and the QRS-T angle, 46 patients (20%) by both Cornell product and the QRS-T angle, and 64 patients (28%) by both the QRS duration and the QRS-T angle, respectively. The QRS-T angle had a 50%, 57% 55% and 51% agreement in correctly classified cases and non-cases with the Sokolow-Lyon Index, Cornell voltage, Cornell Product, and QRS duration, respectively, yielding a Cohen’s kappa (*p*-value) of -0.13 (<0.001), 0.17 (<0.001), 0.13 (0.001), and 0.04 (0.20), respectively. In the combined Healthy-Validation and Imaging-LVH cohort, the QRS-T angle had a higher AUC (AUC 0.99) than all conventional parameters (AUC 0.53–0.91) for detecting LVH (*p*<0.001 for all), see Figure 4. In the Clinical-Consecutive cohort, the AUC for the QRS-T angle (AUC 0.72) did not differ significantly from Cornell voltage (AUC 0.69), Cornell voltage product (AUC 0.71), nor QRS duration (AUC 0.65) (*p*>0.19 for all), see Figure 4.

**Figure 4.**
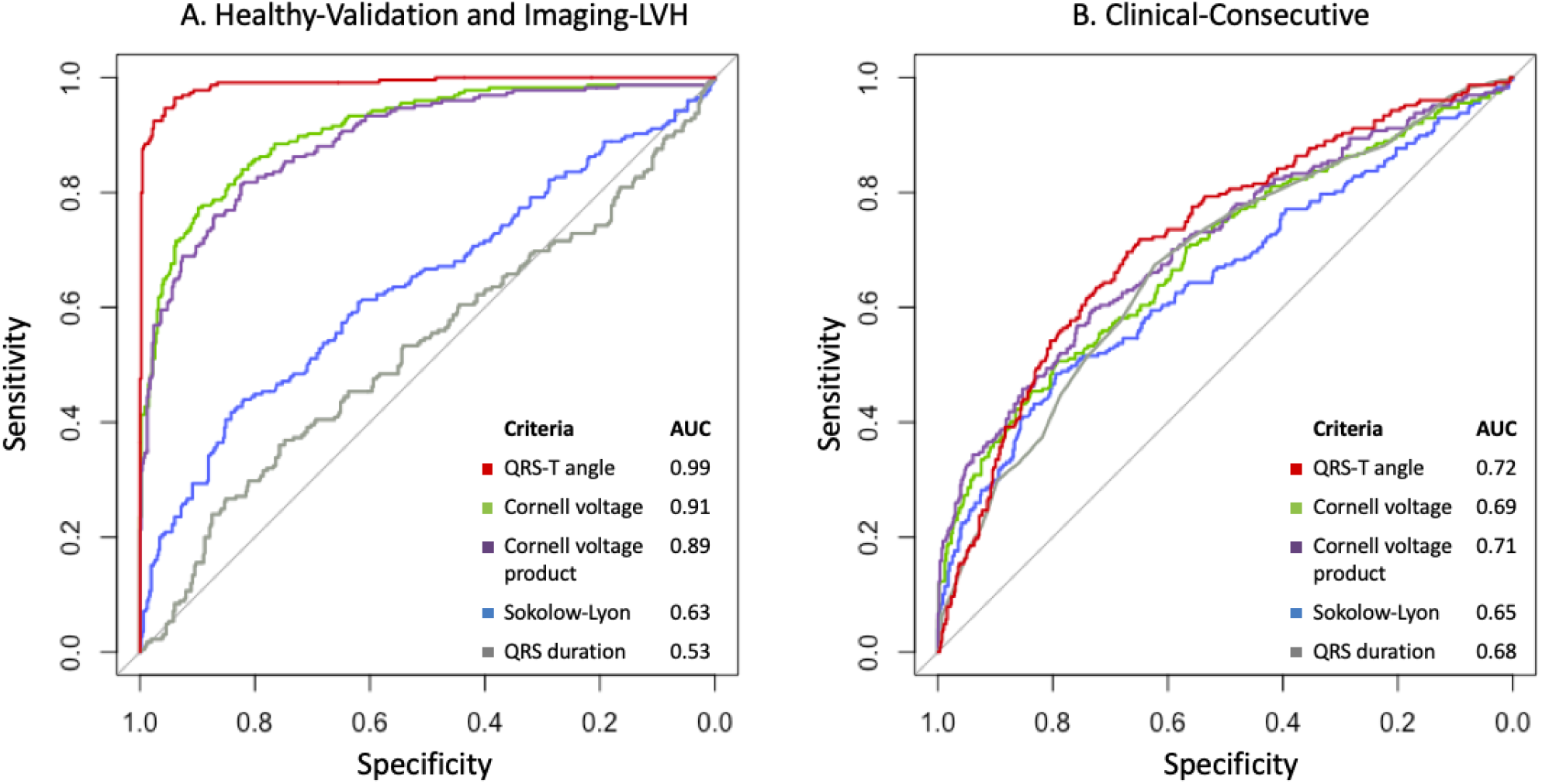
Area under the curve (AUC) for the evaluated criteria for detecting anatomically defined left ventricular hypertrophy. **A.** AUC for the criteria in the combined Healthy-Validation and Imaging-LVH cohorts. **B.** AUC for the criteria in the Clinical-Consecutive cohort.

### The ECG in LVH and outcomes

Per Table 2, univariable Cox analysis showed that the QRS-T angle and Cornell product both had a high univariable association with outcomes (chi-square 22 and 24, respectively, *p*<0.001 for both). Cornell product had a hazard ratio and 95% confidence interval (95% CI) of 2.53 (1.74–3.67), and the QRS-T angle had a hazard ratio of 2.27 (1.62–3.20). Furthermore, an increased QRS-T angle was associated with both components of the combined outcome hospitalization for heart failure and all-cause mortality (chi-square 21 and 10, respectively, *p*<0.001 and *p*<0.01, respectively) with a hazard ratio of 3.94 (2.19–7.10), and 1.86 (1.26 – 2.73), respectively. In multivariable Cox analysis, the Sokolow-Lyon index was excluded due to the absence of an association with outcomes in univariable analysis, and Cornell product was excluded due to both being mathematically related to both QRS duration and Cornell voltage. Multivariable Cox regression showed an association with outcomes for both the QRS-T angle (hazard ratio 1.95 (1.36–2.80), *p*<0.001) and QRS duration (hazard ratio 1.99 (1.42–2.77), p<0.001), but not for Cornell voltage (hazard ratio 1.12 (0.70 – 1.81), *p*=0.63). When comparing the QRS-T angle to co-morbidities, multivariable Cox regression showed an association with outcomes for the QRS-T angle criterion (hazard ratio 1.68 (1.18– 2.38), *p*=0.004), hypertension (hazard ratio 1.96 (1.34–2.87), *p*<0.001), and diabetes mellitus (hazard ratio 2.63 (1.88-3.68), *p*=0.004), but not for smoking status. Figure 5 shows Kaplan-Meier analysis illustrating the poorer event-free survival for patients with an increased QRS-T angle compared to those with a normal QRS-T angle (*p*<0.001). The NRI analysis showed an incremental prognostic value of the QRS-T angle beyond the Cornell product with a NRI of 0.40, 95% CI 0.02–0.66.

**Table 2.** Uni- and multivariable Cox regression analysis for the QRS-T angle, QRS duration, Cornell voltage product, Cornell voltage, and Sokolow-Lyon index in the Clinical-Consecutive cohort for predicting the composite outcome of hospitalization for heart failure and all-cause mortality. Covariates were analyzed as binary variables with cut-offs for left ventricular hypertrophy. See text for details on cut-offs. Cornell product was not included in multivariable analysis due to being mathematically related to QRS duration and Cornell voltage.

| **Covariates** | **Univariable Cox Regression Model** |  |  | **Multivariable Cox Regression Model** |  |  |
| --- | --- | --- | --- | --- | --- | --- |
| | $\chi^2$ | **Hazard ratio**<br>**(95% CI)** | *p*-value | $\chi^2$ | **Hazard ratio**<br>**(95% CI)** | *p*-value |
| **QRS-T angle** | 22 | 2.27 (1.62- 3.20) | <0.001 | 20 | 2.22 (1.56-3.16) | <0.001 |
| **Cornell product** | 24 | 2.53 (1.74- 3.67) | <0.001 | - | - | - |
| **QRS duration** | 18 | 2.02 (1.45- 2.80) | <0.001 | 16 | 1.99 (1.42-2.77) | <0.001 |
| **Cornell voltage** | 7 | 1.81 (1.15- 2.84) | 0.006 | 0.2 | 1.12 (0.70-1.81) | 0.63 |
| **Sokolow-Lyon** | 1 | 1.45 (0.71-2.96) | 0.30 | - | - | - |

**Figure 5.**
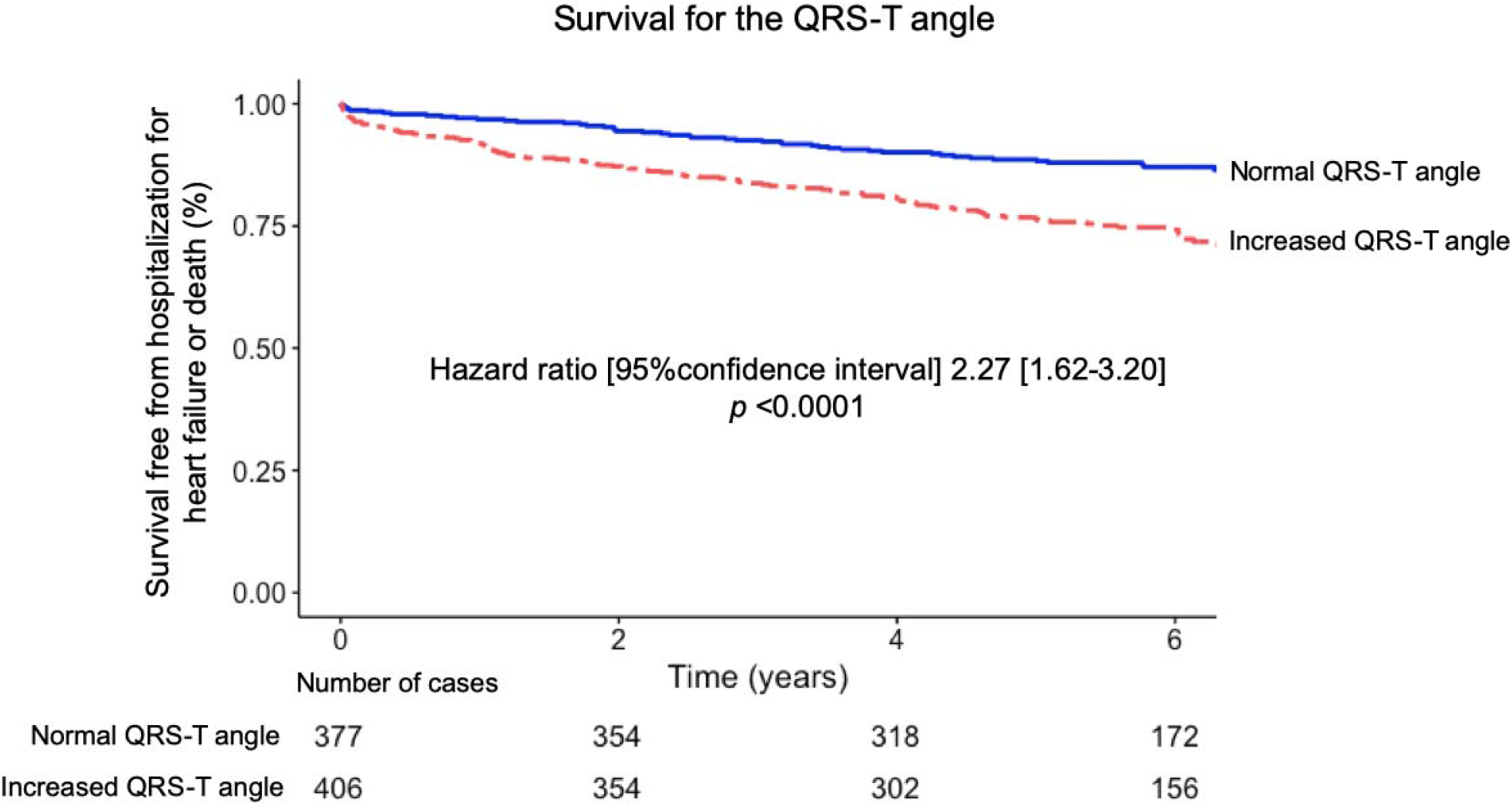
Outcomes for patients in the Clinical-Consecutive cohort according to the presence of a normal or increased derived spatial peaks QRS-T angle, respectively, defined as ≥40° for females and ≥55° for males. An increased QRS-T angle was associated with worse outcomes in the form of survival free from heart failure hospitalization or all-cause death with events in 155 patients over a follow-up period of 5.7 [4.4-6.6] years.

### Relationship between the QRS-T angle and CMR measures

The QRS-T angle correlated with GLS, ECV, LVEDVI, LVMI, infarct size, and non-ischemic scar size (R^2^=0.01–0.25, *p*<0.05 for all). In multivariable analysis, GLS, ECV, LVMI, and infarct size remained associated with the QRS-T angle (model adjusted R^2^=0.28, *p*<0.001).

## Discussion

The major finding of this study was that, compared to existing ECG criteria for LVH, the proposed QRS-T angle criteria substantially improved sensitivity and preserved excellent specificity for identifying moderate or greater anatomical LVH in the Imaging-LVH cohort with at least moderate LVH by imaging. Furthermore, the QRS-T angle also provided important information for risk stratification and outcomes that was independent of that from the Cornell product and co-morbidities, respectively, in multivariable analysis. The current study also found that an increased QRS-T angle is common even among patients referred for clinical CMR who had normal LVMI. Furthermore, abnormal QRS-T angle was exceedingly rare in the Healthy-Validation sub-cohort, which could be expected given that both the Healthy-Derivation and Healthy-Validation groups were randomly selected from the same larger cohort of healthy volunteers. This illustrates that the QRS-T angle identifies an electrical remodeling process often missed by anatomical imaging. This finding is in agreement with the previous finding that anatomical LVH and conventional ECG criteria for LVH possess independent prognostic information (9).

### The spatial peaks QRS-T angle

The spatial peaks QRS-T angle is an established vectorcardiographic measure that can be derived from a standard digital 12-lead ECG. It is defined as the angle in three-dimensional space between the peak magnitude of the vectors for the QRS and T loops, respectively. Similarly to the concordance or discordance between the QRS complex and T waves in a given lead of the conventional ECG, the QRS-T angle reflects the dispersion between the directions of left ventricular depolarization and repolarization (27). In a healthy heart, the angle between the QRS and T loops is small, whereas the angle has been shown to increase as a result of myocardial disease processes (Figures 1–2) (13). On the conventional ECG, repolarization changes such as ST depression and T wave inversion, sometimes referred to as strain, are also known markers of LVH with reported sensitivities and specificities of up to 52% and 95%, respectively (28). Due to a combination of structural, bioelectrical and biochemical changes in LVH (29), the electrical propagation is interrupted, contributing to changes in depolarization and repolarization that usually increase the QRS-T angle. Regardless, the spatial peaks QRS-T angle was more sensitive at detecting LVH compared to conventional ECG criteria that rely upon increased QRS amplitudes or durations. Moreover, by quantifying discordance between depolarization and repolarization, the spatial peaks QRS-T angle also introduces prognostic information independent of QRS amplitude and duration as quantified by Cornell product.

The QRS-T angle has historically been measured by vectorcardiography using acquisition with dedicated lead placement. Vectorcardiography was largely abandoned as a widely used clinical tool in the latter half of the twentieth century due to a combination of technical, interpretation-related, and acquisition standardization-related complexities. However, in today’s healthcare environment, the conventional 12-lead ECG is typically digitally recorded and stored. Consequently, validation of techniques that accurately derive the vectorcardiogram from the digital 12-lead ECG (21) have now made it trivial to automatically and accurately calculate, visualize, and access vectorcardiographic measures including the QRS-T angle.

### Anatomy versus electrocardiography

In the current study, abnormal QRS-T angle was both effectively absent in an independent cohort of healthy volunteers, but present in nearly all patients with imaging-proven, at least moderate, LVH without other predominant cardiac morbidities. The QRS-T angle had a markedly different diagnostic accuracy in the Imaging-LVH and Clinical-Consecutive cohorts, likely due to the presence of intermediate disease in the Clinical-Consecutive cohort, whereas the Imaging-LVH cohort was only based on the presence of moderate or severe LVH. Hence, the presence of intermediate disease, including coronary artery disease without LVH (16), could also increase spatial QRS-T angle values and thus have decreased the diagnostic discriminatory ability in the Clinical-Consecutive cohort. Furthermore, 47% of the Clinical-Consecutive patients underwent a clinical CMR scan due to known or suspected coronary artery disease, potentially reflecting a higher likelihood for presence of disease potentially affecting electrical propagation beyond cardiomyocyte hypertrophy as such. The intermediate diagnostic performance of the QRS-T angle compared to anatomical criteria in the Clinical-Consecutive cohort further underscores the inappropriateness of judging the utility of electrocardiographic criteria using an anatomical arbiter. Due to the known lack of association between electrocardiographic changes and anatomical or structural changes (29), outcomes may be a more appropriate arbiter of diagnostic utility. In fact, the QRS-T angle was associated with prognosis, even beyond that of the conventional criteria with the best performance in both multivariable analysis and NRI analysis. The QRS-T angle had approximately a 50% agreement with the conventional parameters, and a Cohen’s kappa close to 0, indicating that they do not identify the same patients.

### Associations between the QRS-T angle and either CMR or co-morbidities

Compared to patients in the Clinical-Consecutive cohort who had a normal QRS-T angle, those with an increased angle had CMR or clinical demographic characteristics that are associated with a greater severity or burden of cardiovascular disease, with the exception of body mass index or body surface area, and non-ischemic scar size. Despite this, CMR parameters known to convey powerful prognostic information could together only explain approximately one quarter of the QRS-T angle (adjusted R^2^=0.28). This illustrates the complementary information present in the ECG compared to structural and functional measures assessable by imaging. Other groups have also reported a superiority of the QRS-T angle compared to conventional ECG criteria for LVH (11), and that the QRS-T angle is increased in patients with co-morbidities such as hypertension (30) and diabetes (31). However, the current study is unique in that it studied robust sample sizes to both propose sex-specific upper limits of normal for the QRS-T angle, and undertake a comprehensive and definitive approach to evaluation of diagnostic and prognostic performance. Besides the QRS-T angle, the QRS duration was an independent predictor of outcomes, even though patients with complete bundle branch blocks were excluded, while no voltage-related criteria remained associated with outcomes.

### Clinical perspectives

The present study shows a markedly improved diagnostic and prognostic performance of the newly proposed QRS-T angle criterion compared to currently used ECG criteria, suggesting that it provides diagnostic and prognostic information beyond the conventional ECG measures of LVH. The results highlight the difference between the electrocardiographic and anatomical manifestations of disease, and underscore the necessity to define ECG abnormalities as a deviation from ECG normal values in healthy volunteers rather than as defined by anatomical findings. The Journal of Electrocardiology LVH Working Group (10) has proposed that electrical changes associated with LVH may be better defined as LVER as opposed to LVH by ECG. Furthermore, replacing the term LVH with LVER in the evaluation of the ECG has the pedagogical advantage of clarifying the distinction between anatomical and electrical changes associated with remodeling. The current study suggests that an increased QRS-T angle could be used to define LVER, and that the concept of LVER should replace LVH whenever electrical changes are being addressed.

Assessment of the spatial peaks QRS-T angle requires a digitally acquired resting 12-lead ECG and software-based determination. Such solutions may not be available in all clinical settings, but the results of the current study suggest that such implementations may be worth considering. If necessary, the spatial peaks QRS-T angle can also be manually estimated from paper-based 12-lead ECGs using established techniques (32), albeit likely with some compromise in precision and accuracy. Importantly, the presentation of this measure can be easily implemented in a wholly automated fashion into the software of digital ECG machines, and displayed together with conventional parameters and criteria at the time of clinical interpretation.

### Study limitations

Our results are not applicable to patients with QRS duration >120 ms, in whom the QRS-T angle is usually increased, especially in left bundle branch block due to the block itself. Similarly, our results are not applicable to patients with atrial fibrillation or other pathological arrhythmias, whom we excluded due to the known poorer prognosis associated with such arrhythmias (33). The prognostic implications are based on a limited time period with a median of 5.7 years, and no interventions were performed as part of the study. Studies with such interventions and with longer follow up times would be justified. The mean age in the healthy cohorts was younger than those in the Imaging-LVH and Clinical-Consecutive cohorts. As isolated LVH on the ECG is common in younger individuals, the specificity in the healthy cohort would be expected to be lower. Still, the specificity of the QRS-T angle remained high in the Healthy-Validation cohort. Notably, the Healthy-Derivation cohort included an adequate number of subjects over the age of 60 years, and no age differences in the healthy normal range of the QRS-T angle were noted. However, the QRS-T angle could potentially increase with other characteristics that were not evaluated in the current study, and this is a limitation. Furthermore, the 95% upper limit of normal was used to define cut-offs for the QRS-T angle in a large cohort of healthy volunteers, excluding extreme outliers. We did not attempt to distinguish between eccentric hypertrophy from concentric hypertrophy. The QRS-T angle was not hypothesized to be able to perform such a distinction, since both eccentric and concentric hypertrophy can potentially influence the QRS-T angle. Due to the retrospective nature of the study, baseline data were not available for all cohorts. However, this does not affect the prognostic evaluation in the well-characterized Clinical-consecutive cohort. Finally, besides the most common conventional ECG criteria for LVH that we evaluated, other sets of criteria have also been acknowledged by the American Heart Association (34), yet were not specifically studied.

### Conclusions

In conclusion, normal limits for the ECG-based spatial peaks QRS-T angle were defined using a large, healthy population. An increased QRS-T angle rarely occurred in health, was nearly always present in moderate or greater LVH, and was common even in patients without LVH but with cardiac co-morbidities. Increased QRS-T angle was also more accurate than conventional ECG criteria for detecting LVH. Based on the improved diagnostic and prognostic performance of the QRS-T angle compared to conventional ECG criteria for LVH, we propose that QRS-T angle should be investigated when ECGs are evaluated.

## Data Availability

Data can be made available upon reasonable request.

